# Evaluating the impact of the Mellow Babies group-based parenting programme for supporting at-risk mothers in Tajikistan

**DOI:** 10.1101/2024.04.24.24306290

**Authors:** Natalie Duncan, Ailsa Jones, Rachel Tainsh, Nazira Muhamedjonova, Raquib Ibrahim, Angus MacBeth

## Abstract

Parental mental health has a long-lasting impact on developmental outcomes for infants and children through its impact on the family environment. Targeted parenting interventions should address both parental health and parenting skills. However, data is limited on how interventions perform in Central Asian populations.

Using routine evaluation data from n=194 participants, we modelled the effectiveness of the Mellow Babies (MB) programmes, delivered to mothers from Tajikistan and their children as part of a community support approach. Pre-post intervention changes were measured on depression, anxiety, parenting stress, quality of life, and child behaviour outcomes. Demographics were modelled as covariates.

Participation in MB was associated with improvements in maternal mental health, parenting stress, quality of life and child behaviour. Demographic factors moderated interactions between pre and post intervention outcomes, including urban/rural differences, parental employment, marital status and child disability status. Our findings suggest that MB is acceptable, effective and potentially scalable as a parenting intervention in Tajikistan. Future replication using implementation designs and replication of MB in other global settings is merited.

---

Multiple risk factors have been linked to poor developmental outcomes for children including poor maternal mental health, poverty, and reduced access to healthcare(1, 2, 3). Parents’ mental health and well-being have a formative and long-lasting impact on developmental outcomes for infants and children. Parental mental health affects young children through its impact on parent-infant interaction and the family environment (4).

Maternal depression occurs in around 13–15% of women(5), and chronicity in maternal depression across more than one timepoints is associated with an elevated risk of multiple suboptimal developmental outcomes in children, including elevated rates of externalizing and internalizing problems, poorer achievement of overall developmental milestones in childhood(6). Persistent maternal depression in the first 12-months postpartum has long-range effects on behavioural, social, emotional and educational outcomes such as educational attainment at 16years of age (4, 7, 8). This may also extend to mothers who experienced sub-clinical depressive symptoms (9).

Further, maternal anxiety during pregnancy and increased child difficulties such as emotional symptoms, conduct problems and hyperactivity at age 10 (10).

However, these risks may themselves be nested within broader social determinants of health and childhood experiences of poverty have been consistently linked to poor developmental and mental health outcomes(11). Exposure to poverty during childhood increases risk of internalised symptoms, impacting upon psychological wellbeing, and higher instances of externalising behaviours (12). Maternal mental health may also act as a mediating factor between social economic status (SES) and developmental outcomes. Child mental health and cognitive ability may mediated by maternal mental health, particularly prior to 1 year postnatal(13). This study highlights the importance of positive maternal mental health as a protective factor for child wellbeing and learning, particularly within the first year of the child’s life. Further, children of mothers with self-reported positive mental health, support from others, and lower levels of distress displayed better mental health than their peers (14); and associations have been reported between cognitive delay and food insecurity, mediated by maternal mental health(15).

## Early intervention parenting programmes

From a public health perspective the above evidence highlights the importance of targeting interventions towards mothers in low-income families (14, 15) and that interventions should aim to promote positive childhood experiences (16). Factors including timing and severity of adversity may also be important methodological considerations with evidence suggesting that severe maternal deprivation experienced between 0 – 2 months had a significant impact on functioning in middle childhood and these effects were greater that at earlier developmental stages (17). The study provides evidence and a rationale for engaging in early intervention and prevention of childhood trauma.

Meta-analytic studies indicated that parenting interventions (such as Triple P, Incredible Years and Mellow Parenting (MP)) can have a positive impact on children’s behaviour alongside improving parenting skills and parental wellbeing (18, 19). However, there are challenges with the existing evidence base for these interventions including recruitment biases towards including middle-class families in most of the research, raising concerns about the exclusion of families from low socioeconomic backgrounds, over-reliance in the evidence base on data from high income countries (19)and challenges in establishing the independence of evaluation frameworks from intervention delivery (20).

Although the evidence base for delivery of parenting programmes in low and middle income countries is growing (21) these generally focus on the impact on parenting approaches and child outcomes, with relatively less focus on the impact on caregiver mental health. There are also challenges remaining around how these evaluations work with contextual factors such as poverty, adversity and cultural sensitivity.

There is a growing body of evidence around the application of parenting programmes that use combined targeting of parenting, child behaviour and parental health outcomes. A UK evaluation of MP reported improvements in maternal mental health, parenting confidence, and child conduct problems after participation in the programme(22). Further, most participants in these programmes self-identified as unemployed and presenting with other population level vulnerabilities. A subsequent evaluation of MP reported similar outcomes in terms of wellbeing and confidence for both mothers and fathers who participated in the intervention (23). However, there is limited recent evaluation evidence of the effectiveness of Mellow Parenting in LMIC settings(24).

## Tajikistan context

The current study evaluated the delivery of Mellow Babies (a version of Mellow Parenting adapted for infants) in the Central Asia nation of Tajikistan. Mellow Parenting has been adapted to be delivered in non-high resource settings such as Central and Eastern Europe and the Commonwealth of Independent States (CEE/CIS), where health policy has moved to cease the practice of end institutional care rearing for children under the age of 3 years, replacing this with national social protection systems supporting children to live within a family environment. Mellow Parenting groups were introduced to Tajikistan in 2010 in the context of a gatekeeping and reintegration approach to develop family support alternatives and prevent babies at risk entering institutional care settings such as Baby Homes (25). The Putting Families First Project also built on previous work to develop family support and early intervention alternatives to institutional care and included Mellow Parenting Programmes as part of a package of support for families in the community (26).

Tajikistan has seen a steady decrease in poverty rates over the past decade but continues to face economic challenges following COVID-19 and the recent conflict in Ukraine (27). High underemployment within Tajikistan has been linked to high levels of Migration to neighbouring countries such as Russia, which in turn leads to increased vulnerability for “left-behind” women and children (28).

Research suggests high prevalence rates for depression (26%), anxiety (17%), and PTSD (17%) amongst Tajik women (29). Further, the high levels of migration mean that Tajik wives are vulnerable to being separated from their husbands, having little-to-no financial or social support, and often left dependent on their in-laws (30). Evidence suggests that Tajik women are at high risk of domestic violence at the hands of their husband/in-laws and at least 23% of married women have experienced domestic violence (31). Violence against women and girls (VAWG) is a major problem in Tajikistan, driven by conservative gender norms, the culturally ascribed position of young women, and poverty, although there is evidence for the effectiveness of violence reduction programmes (30). Tajik women also have elevated levels of stress and poor living conditions, with suicide rates increasing by 176% from 2008 – 2010, and suicide completion and attempt statistics for Tajikistan were reported as three suicides for every one attempt, in contrast to the global statistics one suicide for every 10 – 20 attempts (32). As childhood experiences of poverty, poor maternal health and limited access to healthcare all increase the risk of poor child outcomes, targeted early interventions for at-risk families in Tajikistan may be one way to mitigate the impact of these social and demographic risk factors.

## Current Project

The current study analysed pre– and post-data collected from mothers who participated in the Mellow Babies intervention, targeting families of babies aged 0 – 18 months. We sought to evaluate associations between participation in a Mellow Babies group and maternal mental health outcomes for Tajik mothers. The secondary aim was to evaluate evidence for associated improvements in parenting stress, quality of life, and child behaviour following participation in a Mellow Babies group. Finally, we aimed to identify baseline predictors of outcomes in the sample. It was hypothesised that participation in a group would be associated with improved maternal mental health, and reduced parenting stress but that sociodemographic factors would affect the degree of improvement on outcomes.

## Materials and Methods

### Design

An uncontrolled retrospective cohort design was used to assess pre-post intervention changes on the primary and secondary outcomes. Data were available from participants who took part in Mellow Babies interventions in Tajikistan between 2016 – 2020 and were collected as part of participation in the intervention. Participants were aware that anonymised data would be used for subsequent evaluation of the programme.

### Participants and eligibility

To be eligible for participation in the group mothers had to have at least one child under the age of 18 months with whom they had frequent contact. It was a requirement that their children participated in Mellow Babies groups.

The total sample was n=195. The mean age of participants was 33.20 years of age (SD=7.52), and the mean age of babies was 5.52 months (SD=3.09). All participants attended 12 or more sessions of the programme. There were 194 participants who completed at least one outcome measure at baseline and post intervention. Figure 1 details the flow of participant data through the study.

**Figure 1:**
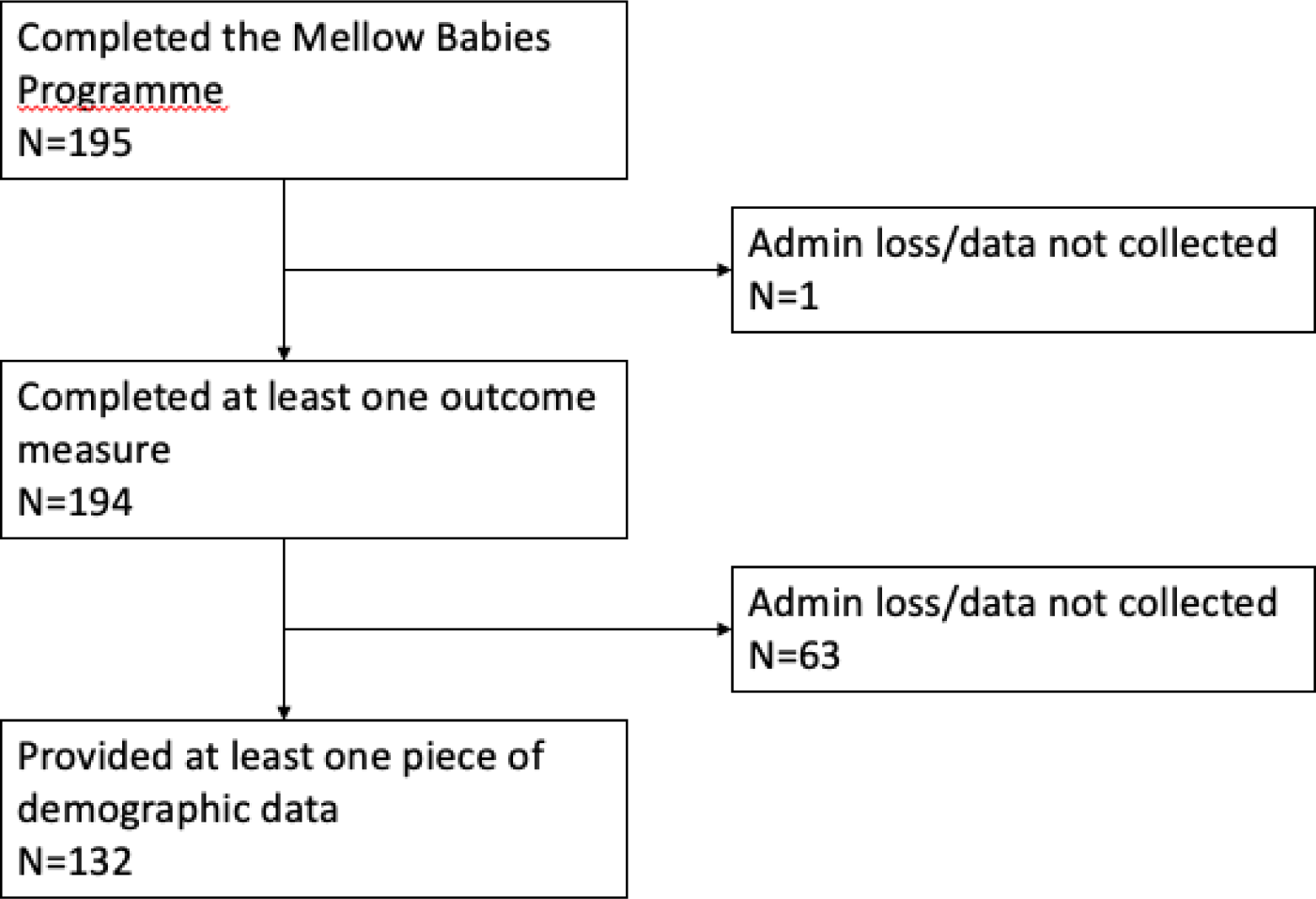
Flow diagram of participant data in study.

### Measures Depression and Anxiety Stress Scale 21 (DASS-21)

The DASS-21 is a 21-item self-report questionnaire measuring depression, anxiety, and stress. Statements are presented on a 4-point Likert scale based on frequency of symptom. The DASS-21has high internal consistency overall and across each construct (Depression α = .88; Anxiety α = .82; Stress α = .90; Overall α =.93; (33)). Cross-cultural studies have also found evidence for reliability and validity when administering the DASS-21 across different settings (34).

### Parenting Daily Hassle Scale (PDHS)

The PDHS is a 20-item self-report questionnaire measuring daily parenting stress based on how often the parenting challenges occur (on a 4-point Likert scale) and their intensity (5-point Likert scale). The frequency is scored out of 80 and the intensity score is out of 100. Both measures have acceptable internal consistency (frequency α = .81 and intensity α = .91; (35). The PDHS has two subscales measuring challenging behaviour (single item factor loading ranged from α = .31 – .81) and parenting tasks (single item factor loading ranged from α = .40 – .78) (35).

### Quality of Life, Enjoyment and Satisfaction Scale (QoLESS)

The QoLESS is a 16-item self-report questionnaire measuring overall quality of life in terms of satisfaction and enjoyment in areas of health, work, relationships, and overall wellbeing. Statements are presented on a 5-point Likert scale ranging from “very poor” to “very good”. It has high internal consistency (α = .90) and test-retest reliability (α = .93;(36)).

### Richman’s Behaviour Checklist (RBCL)

The RBCL is a 21-item parent-reported questionnaire that measures child behaviour problems. Each item presents three statements and asks the parent to put an “x” in the box corresponding to the statement that is most true to their child’s behaviour. RBCL was reported to have high test re-test reliability (r=.81;(36, 37)).

### Procedure

Mellow Babies groups were delivered as part of the Putting Families First Project. All participants lived in Tajikistan and were identified based on their previous involvement with the organisation. Mothers were asked to provide consent to participate in the evaluation before data was collected. Over the study period 16 separate Mellow Babies groups were delivered. Each participant took part in one group with the numbers of participants in each group ranging from five to twelve. Fifteen of the 16 groups met face-to-face and one group of ten met online. Participants consented to taking part in the Mellow Babies Programme. Pre-intervention measures were completed before the first session.

The groups took place over 14 weeks, one session per week, and were facilitated by two trained Mellow Parenting facilitators. Each session lasted 4.5 hours and was split into morning, lunch, and afternoon activities. The morning activity consisted of parents reflecting childhood and the ways in which that may have impacted their current parenting approach. The second part of the session was lunch, during this time parents spent time with their child on a joint activity. The aim of this was to strengthen the parent-child relationship. The third part of the day was the workshop which focused on parents’ videos of interactions with their child. This activity provided an opportunity for participants to receive supportive feedback from facilitators and fellow parents. The child groups took place alongside parent sessions, allowing parents to participate in morning and afternoon activities. All post-intervention measures were collected at either on session 13, 14 or at a home visit after last session.

## Data analysis and approvals

Ethical approval for the data analysis was received from the University of Edinburgh Health in Social Science Ethics Committee. Associations between participation in MB and outcomes were analysed using paired-samples t-tests. Where data were non-parametric the Wilcoxon signed rank test was used. Pearson’s correlations were conducted to test for associations between baseline demographics and scores on pre-intervention measures. Finally, one-way ANCOVAs were carried out to measure the effect of categorical demographics on outcomes, assessing interactions between pre and post scores controlling for the following baseline demographics: residence, education, employment, child disability, and marital status. Effect sizes were reported as Cohen’s d for pre-post comparisons, and η ^2^ for ANCOVAs.

## Results

### Demographics

Baseline demographics are displayed in Table 1. Based on the available data, the majority of participants were of Tajik nationality, living in an urban setting. Most participants reported either being married or living with their partner. Almost two thirds of participants were unemployed. The majority of participants had education to secondary school level or above. With regard to children, the majority of participants identified their child as having a disability. Where recorded attendance rates were very high with all participants attending at least 12 sessions out of 14, and 83.8% of participants attending all 14 sessions (based on 79.8% of all pre-post evaluation data).

**Table 1.** Participant demographics and engagement with programme.

| Study variable | Frequency (%) |
| --- | --- |
| **Nationality (N=132)** |  |
| Tajik | 121 (91.7%) |
| Uzbek | 8 (6.1%) |
| Other | 3 (2.2%) |
| **Residence (N=132)** |  |
| Urban | 110 (83.3%) |
| Rural | 22 (16.7%) |
| **Marital Status (N=127)** |  |
| Married/living together | 106 (83.5%) |
| Single/Divorced/separated/widowed | 21 (16.5%) |
| **Employment (N=108)** |  |
| Unemployed | 72 (66.7%) |
| Employed | 36 (33.3%) |
| **Education (N=108)** |  |
| Primary | 11 (10.2%) |
| Secondary | 71 (65.7%) |
| Further/higher | 26 (24.1%) |
| **Child disability (N=130)** |  |
| No | 40 (30.8%) |
| Yes | 90 (68.2%) |
| **Participant disability (N=187)** |  |
| No | 86 (98.9%) |
| Yes | 1 (1.1%) |
| **Number of sessions attended (n=154)** |  |
| 12 | 7 (4.5%) |
| 13 | 18 (11.7%) |
| 14 (all sessions) | 129 (83.8%) |

## Mental health, parenting and child outcomes

Pre-post intervention comparisons (Table 2) reported significant pre-post intervention reductions on maternal DASS total scores (t(193) = 13.69, p<.001), Depression (t(193) = 13.29, p<.001), Anxiety (Z = –9.39, p<.001) and Stress (Z = –9.72, p<0.001) subscales. For parenting hassles, PDHS Intensity and Frequency significantly reduced from pre to post intervention (t(181) = 14.30, p<.001; Z = –9.41, p<0.001), as did the Parenting Tasks and Challenging Behaviour subscales (t(181) = 11.38, p<.001; t(181) = 12.88, p<.001). Finally, QoLESS scores indicated a significant improvement in self-reported quality of life across the intervention (t(173) = –13.63, p<.001, d = 1.03). There was also a significant decrease in child behaviour (RBCL Z = –8.98 p<.001). Cohen’s *d* indicated large effect sizes across all tests. Due to the number of t-tests carried out, the p value was adjusted using an alpha correction by calculating p value/number of t-tests resulting in a corrected p-value of p<.005. All tests remained significant after correction.

**Table 2.** Pre and post mean scores for outcome measures.

| Outcome measure | Pre mean (SD) | Post mean (SD) | Cohen's d (95% CI) |
| --- | --- | --- | --- |
| **DASS Total (n=194)** | 81.48 (28.79) | 47.24 (26.07) ** | 0.98 (0.81 to 1.15) |
| **Depression (n=194)** | 13.01 (5.58) | 7.09 (4.86) ** | 0.95 (0.78 to 1.12) |
| **Anxiety (n=194)** | 13.01 (5.24) | 7.64 (4.48) ** | 0.86 (0.69 to 1.02) |
| **Stress (n=194)** | 14.73 (4.55) | 8.88 (4.44) ** | 0.96 (0.79 to 1.14) |
| **PDHS Frequency (n=182)** | 50.47 (13.59) | 39.38 (12.75) ** | 0.86 (0.70 to 1.03) |
| **PDHS Intensity (n=182)** | 63.42 (19.57) | 44.96 (17.85) ** | 1.06 (0.88 to 1.24) |
| **PDHS Parenting tasks (n=182)** | 25.44 (8.09) | 18.84 (7.81) ** | 0.84 (0.67 to 1.01) |
| **PDHS Challenging behaviour (n=182)** | 22.93 (7.11) | 16.61 (6.55) ** | 0.95 (0.78 to 1.13) |
| **QoLESS (n=174)** | 40.17 (10.00) | 50.40 (8.69) ** | 1.03 (0.85 to 1.22) |
| **RPBC (n=193)** | 4.63 (4.38) | 1.99 (2.64) ** | 0.76 (0.60 to 0.92) |

Notes: \*\* p<0.01; DASS = Depression, Anxiety and Stress Scale; PDHS = Parenting Daily Hassles Scale; QoLESS = Quality of Life, Enjoyment and Satisfaction Scale; RBCL = Richman's Behaviour Checklist; 95% CI = 95% Confidence Interval

## Relationships between baseline demographics and outcomes

There were significant correlations between the child’s age and both baseline PDHS Intensity (r(127) = –.208 p = .018) and Challenging Behaviour (r(129) = –.295 p = .001). This indicated that parents of younger children reported increased difficulties on these scales. Further, there were negative correlations between child age and PDHS Frequency (r(129) = –.243, p = 0.005) and RBCL Child behaviour scores (r(129) = –.308, p<.001. This indicated that parents of younger children also reported increased difficulties on these scales. Child age was not significantly correlated with maternal DASS score or Quality of life. Participant age was not significantly correlated with baseline measures.

## Interactions between baseline demographics and outcomes

ANCOVA analyses were carried out to assess interactions between baseline scores and the baseline demographics (residence, education, employment, child disability, and marital status) on outcome (see Table 3).

**Table 3.**
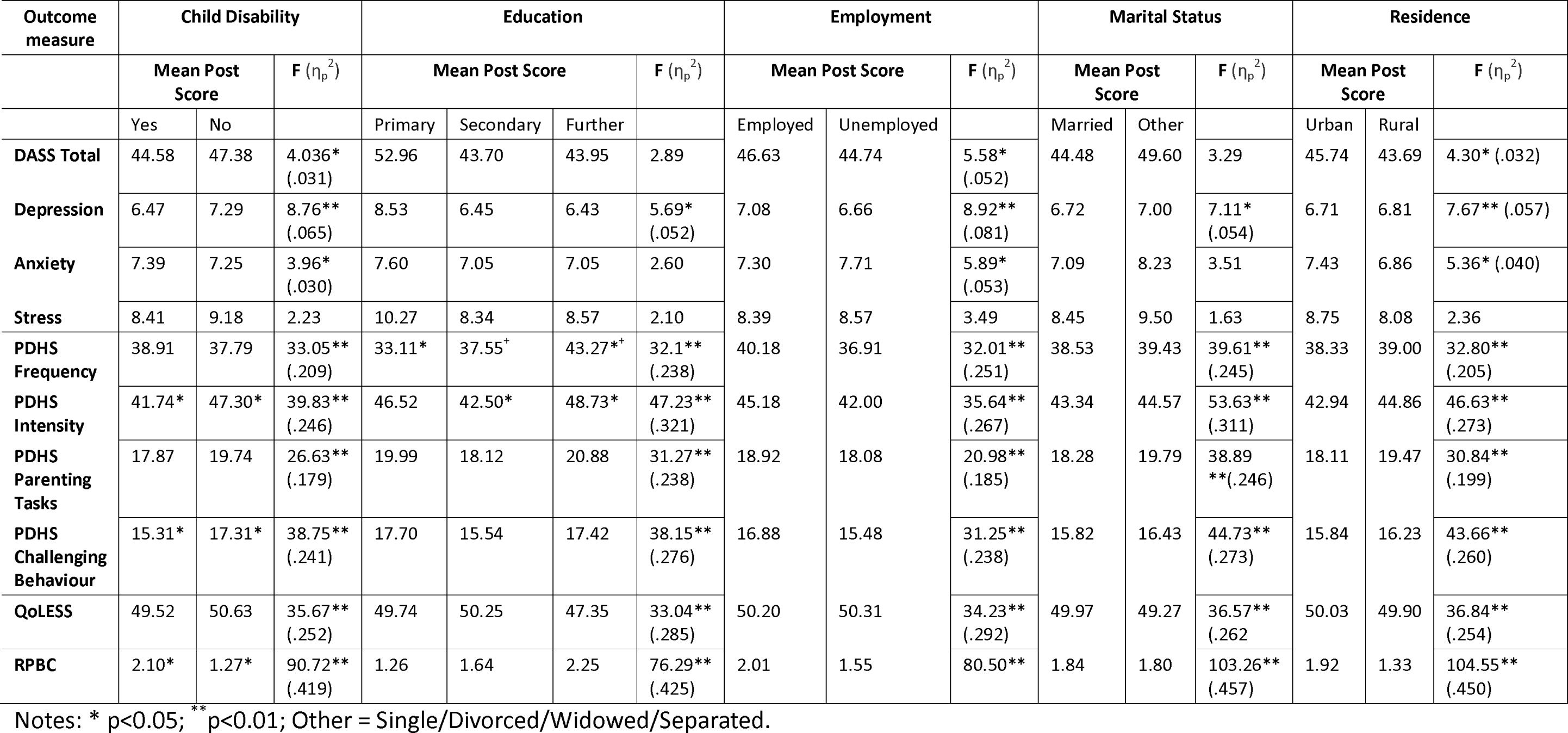
Interactions between basic demographics and post-treatment outcomes.

### Child disability status

There was a significant interaction between pre-intervention score and disability status for post-intervention DASS total scores (p = .047, 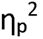 = .031), Depression ( p=.004 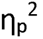 =.065) and Anxiety (p=.049, 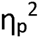 =.030); for PDHS Frequency (p<.001, 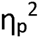 =.209) and Intensity (p<.001, 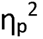 =.246), Parenting Tasks (p<.001, 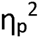 =.179), Challenging Behaviour (p<.001, 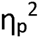 =.241), Quality of Life (p<.001, 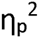 =.252) and child behaviour (p<.001 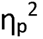 =.419). These indicated significantly lower self-reported post-intervention outcomes for parents of disabled children compared to parents whose children were not disabled, across DASS total, depression, hassle intensity, parenting tasks and challenging behaviour. In contrast, parents of children without a disability self-reported lower anxiety, hassle frequency, and problem behaviour and higher levels of quality-of-life post-intervention.

### Highest education level

Significant interactions were also evident between the pre-intervention score and highest education level for post-intervention Depression F(p=.019 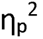 =.052), PDHS Frequency (p<.001 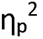 =.238), Intensity (p<.001 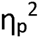 =.321), Parenting Tasks (p<.001 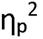 =.238), Challenging Behaviour (p<.001 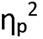 =.276), Quality of Life (p<.001 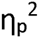 =.285) and child behaviour (p<.001 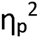 =.425) outcomes. Participants with primary-level education reported the highest rates of depression on the DASS post-intervention. Those educated to further-education level reported the highest level of difficulties across the parenting scales, the challenging and problem behaviour scales and lower quality of life post-intervention compared to other groups. There were no significant interactions between highest education level and pre-intervention scores for DASS Total, Anxiety or Stress.

### Residence

There were significant interactions between the pre-intervention score and residence for post-intervention DASS total F(p=.040, 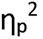 =.032), Depression (p=.006, 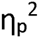 =.057), Anxiety (p=.022, 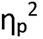 =.040), PDHS Frequency (p<.001, 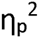 =.205), Intensity (p<.001, 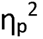 =.273), Parenting Tasks (p<.001, 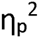 =.199), Challenging Behaviour (p<.001, 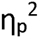 =.260), Quality of Life (p<.001, 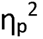=.254), and child behaviour (p<.001, 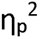 =.450). Those living in urban areas reported significantly lower levels post-intervention depression, hassle frequency and intensity, parenting tasks, challenging behaviour, and higher quality-of-life compared to rural participants. Conversely, individuals in rural areas scored significantly lower than urban participants on post-intervention DASS total and Anxiety scales and reported lower levels of child problem behaviour. There was no significant interaction for DASS Stress outcomes.

### Employment status

For each of the following outcomes there was a significant interaction between the pre-intervention score and employment status on post-intervention outcomes: DASS total (p=.020, 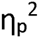 =.052), Depression p=.004 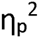 =.081), Anxiety (p=.018 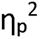 =.053), PDHS Frequency (p<.001 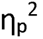 =.251), Intensity (p<.001, 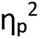 =.267), Parenting Tasks (p<.001 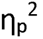 =.185) and Challenging Behaviour (p<.001, 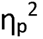 =.238) Quality of life (p<.001, 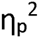 =.292) and Child behaviour (p<.001 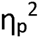 =.425). This indicated that participants in employment reported higher rates of mental health symptoms, depression, parenting hassles and challenging/problem child behaviour compared with unemployed participants. Employed participants also reported slightly lower quality of life scores post-intervention. However, post-intervention anxiety scores were higher in unemployed participants than employed participants. There was no significant interaction between employment status and pre-intervention scores on Stress outcomes.

### Marital status

For each of the following outcomes there was a significant interaction between pre-intervention score and marital status on post-intervention scores: DASS Depression (p=.009, 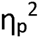 =.054), PDHS Frequency (p<.001, 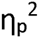 =.245), Intensity (p<.001, 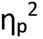 =.311), Parenting Tasks (p<.001, 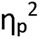 =.246), Challenging Behaviour (p<.001, 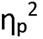 =.273), Quality of life (p<.001, 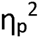 =.262) and child behaviour (p<.001, 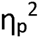 =.457). Unmarried participants reported higher rates of post-intervention depression, parenting hassles, and challenging behaviour compared with married participants. However, post-intervention quality of life was slightly lower for unmarried participants.

## Discussion

Our results represent the largest evaluation to date of a parent and infant mental health interventions in Tajikistan to date. Our findings suggest participation in Mellow Parenting was associated with significant post-intervention improvements in self-reported parental mental health capacity to manage parenting stress, reduced problematic child behaviour, and improved quality of life at the end of the intervention; with all changes consistent with large effect sizes. These findings are consistent with existing research into the effectiveness of MP (22, 23), but extend the evidence base into middle resource settings, and with consistently larger effect sizes. The mean age of children in the sample was around 5, suggesting that MP is a potentially effective intervention in the perinatal period. Further, MP was acceptable and feasible to deliver in the Tajik context, with over 80% of participants attending all 14 group sessions, representing higher engagement than UK samples(38). Pre-intervention data suggested mean levels of depression and anxiety in the sample consistent with mild common mental disorder, which had dropped to minimal levels post-intervention.

The current study also reported improvements in child behaviour and parenting hassles, suggesting that participation in MP is associated with broad changes in both maternal mental health and parenting outcomes and may ultimately improve child-related outcomes. This is consistent with network mapping evidence from Tajikistan, indicating that availability of psychological support was linked to more positive relationships between caregivers and children(39).

Further, demographic data indicated a broad range of markers indicative of increased vulnerability within the sample including high levels of unemployment, child disability and education ending at secondary school level. Our demographic data also suggested that vulnerability and risk in mother-infant dyads may be nested within demographic vulnerabilities, consistent with the observation that maternal mental health chronicity emerges within the context of social determinants (6). For instance, those participants who were not currently married showed higher levels of mental health symptoms post-intervention. Participants in rural areas reported higher levels of depression and parenting challenges at post-intervention than those in urban areas, although rural mothers also reported lower anxiety. In addition, employment was associated with higher levels of distress and parenting challenges, perhaps signalling stress emerging from pressure to maintain livelihoods. These differential findings for specific demographic factors highlight the challenges of matching intervention delivery to need.

Given the Tajik context, these outcomes could be interpreted in terms of levels of support available to mothers. Tajikistan has moved towards developing family support and child centres which may be accessed by parents of disabled children (40). Within Tajik culture, it is also common for husbands to migrate for employment to provide and for women to live with extended family (41). Therefore, mothers who identified as “unemployed” may be receiving support from in-laws. Consequently, future evaluations should take into account the role of cultural context as both a baseline factor and a process factor in the delivery of the intervention.

In addition, pre-post intervention change on most of our secondary outcomes (parenting, child behaviour, and quality of life) were subject to significant interactions with baseline demographic characteristics. More positive parenting outcomes were associated with demographics such as having a child with a disability, living in an urban area, being unemployed and being married, and lower levels of post-intervention child behaviour problems were associated with baseline unemployment, and marriage. With the exception of unemployment, each of these interactions could represent the impact of access to wider or more organised support mechanisms (e.g. community, health and social care availability). Further, rural location was also associated with lower child behaviour problems, which could also represent availability of supports in the wider family and community. For disabled children, parents reported less challenging behaviour on the PDHS but increased behaviour problems on the RBCL. Over recent years there have been increases in the level of support available to Tajik mothers, particularly those with additional needs, including the family support centres and living with in laws (40, 41). However, more research is needed to unpick the nature of interactions between intervention and demographic factors.

In addition, we identified significant correlations between child age and parenting stress suggesting that mothers of younger children identified elevated levels of parenting stress and higher levels of behaviour problems. This could fit with the relevance of sensitive periods in child development . For instance, mothers adjusting to the parental role, particularly in the context of additional needs or existing vulnerabilities could experience increased levels of stress in the early months post-partum. Equally, mothers may be more aware of challenging/problem behaviours and accordingly more likely to report them.

## Strengths of the current research

The present study builds on existing research on the effectiveness of Mellow Parenting (18, 23, 38) in improving infant and parental mental health, extending this work to a global setting, in a middle resource nation. This adds to the cross-cultural evidence base for MP. Further, the outcome measures used to collect data were all standardised questionnaires, providing high levels of reliability and enabling comparison with other MP programmes and other parenting programmes in general. In addition, the high level of engagement indicates that MP is acceptable to, and well-tolerated by Tajik participants, indicating that there is a basis for scaling up both intervention and evaluation of MP in the country. Furthermore, delivery of the intervention occurred in different sites within Tajikistan, Khuajand, Panjakent and Dushanbe, within community settings, and offered an example of positive NGO government partnership working.

That said, we acknowledge several limitations. Due to the “real world” design of the research, there was no control group, limiting comparisons with families who did not receive an intervention. Therefore, it was not clear if improvements entirely were due to the programme or non-specific factors such as time. Further, given that the data was collected as part of routine delivery of MP in Tajikistan, demographic data was missing for many participants. Although data was collected using standardised measures, two measures (RBCL and PDHS) were administered to children outwith the intended age-group. In addition, only total scores of each measure were recorded. Therefore, it was not possible to record item-level validity. In addition, given that participants’ children in the current study were 18 months and younger, the findings for parenting stress and child outcomes should be interpreted with caution. Future research should aim to use outcome measures which have been standardised for the age groups participating in the study (22). Finally, all outcomes used in the present research relied on self-report measures, reported by mothers who participated in the Mellow Babies group. Future research should therefore aim to collect observational data to accompany self-reported outcomes.

In summary, our evaluation identified that, for Tajik mothers, participation in the Mellow Babies programme was associated with improvements in parental mental health, parenting, child, and quality-of-life outcomes. Future research is required to evaluate MP using controlled trials or implementation designs, as well as examining the long-term impact of participating in Mellow Parenting. Further, our findings suggest that there is sufficient infrastructure in Tajikistan to support this work (42).

## Data Availability

All data produced in the present study are available upon reasonable request to the authors

